# Changes in English NHS outpatient activity during the early Covid-19 period

**DOI:** 10.1101/2021.04.28.21256176

**Authors:** Jessica Morris, Theo Georghiou, John Appleby

## Abstract

**Objective:** To describe changes in NHS outpatient activity connected to the Covid-19 pandemic

**Design:** Nationwide population-based retrospective study

**Setting:** England, UK, 31 December 2018 to 25 October 2020

**Data source:** Outpatient Hospital Episode Statistics data

**Results:** Between early March and late October 2020, there was a total reduction of 16.6 million outpatient attendances compared to the same period in 2019, equivalent to a 27% decline. The largest weekly drop of 48% relative to 2019 occurred the week beginning 30 March. Activity recovered more slowly than it fell, and by the end of the study period remained 16% lower than the equivalent week in 2019. Changes in patterns of attendances were broadly similar across most patient characteristic groups. There was a substantial increase in the proportion of attendances taking place remotely, peaking at more than one in three during April and May 2020. Differences were observed in trends of remote consultations between age and sex categories, ethnic groups, and proxy deprivation levels. There was also substantial variation in overall activity and use of remote consultations by clinical specialty.

**Conclusions:** The large increase in remote outpatient consultations during the early Covid-19 period, variations in remote care use by specialty as well as proxy deprivation and ethnic groups all suggest a need to evaluate the impact of these changes particularly in light of national policy to encourage greater use of remote consultations.

**SUMMARY BOX:** *What is already known on this topic:*

- Numbers of outpatient attendances in England have increased substantially over recent years.
- Historically, the vast majority of attendances have been face-to-face, with remote consultations accounting for ∼4% of all attendances.
- The emergence of the Covid-19 pandemic in 2020 had a large impact on outpatient services in English hospitals, and on health services more generally in England and around the world.

*What this study adds:*

- There were significant differences by clinical specialty in changes to patterns of attendances and remote consultations.
- Changes in the volume of outpatient attendances as a result of responses to the pandemic were broadly similar across patient groups.
- However, there were marked differences across patient groups in trends in remote consultations.

## INTRODUCTION

In England, the total number of outpatient appointments has increased by two thirds since 2008/09 - to 125 million a year^1^ – and outpatient services currently account for approximately seven per cent of the National Health Service (NHS) budget.^2^ This is the largest increase in activity of any hospital service, and as a result, long wait times, delayed appointments, and rushed consultations have become more common.^3^ Traditional outpatient service models have relied on face-to-face consultations, which can require repeat hospital visits that often prolong uncertainty and waste patient and staff time.^3^ As part of its outpatient redesign programme, NHS England is looking to avoid one third of face-to-face outpatient attendances by 2024. It claims that this would save the NHS in England an estimated £1.1 billion a year (also saving patients 30 million visits to hospital) by streamlining service delivery through expanded technology at each stage of the pathway.^4^

In spring 2020, soon after the first full year of NHS England’s ten-year programme, the Covid-19 pandemic brought huge disruption to people’s lives, to the economy, and to the nation’s health services. An early priority of the NHS was to ensure that there was available bed capacity to deal with Covid-19 patients. In mid-March 2020, NHS England recommended that hospital trusts should postpone all non-urgent elective operations from mid-April 2020, with local discretion to move quicker than this target.^5^ In wider society, a national lockdown followed on 23 March 2020.

The impact on English NHS outpatient services was significant (just as it was on health services in many other countries).^6^ Total outpatient attendances fell by 48%, from an average of around 8.1 million per month pre-pandemic to 4.2 million in April 2020. By September, attendances had recovered to around 90% of their level in September 2019. The impact of another wave of Covid-19 during winter was evident with attendances in January 2021 falling back to 73% compared to the number in January 2020.^7^ The disruption to outpatient and other NHS services has raised concerns about negative consequences for patient care, including increased waiting times, impact on patients’ health and lost service productivity. However, there are also questions about whether the disruption has led to some positive changes in how the NHS delivers its services – such as greater use of remote consultations, reductions in attendances of perhaps marginal health value, and other innovations.

### Aims

This study is one component of a larger project by the National Institute for Health Research (NIHR) funded Rapid Service Evaluation Team (RSET) which focuses on identifying promising innovations in outpatient services for evaluation.^8^ The aim of this analysis is to describe changes in NHS outpatient activity in England including the use of remote consultations over an initial Covid-19 period (up to late October 2020), in order to inform future work to identify and evaluate innovations in how outpatient services are delivered. We intend to update this analysis periodically.

## METHODS

### Data

We extracted data on all activity recorded in Hospital Episode Statistics (HES) Outpatient dataset between 30 December 2019 and 25 October 2020 inclusive, and for an equivalent period the prior year (31 December 2018 to 27 October 2019). These dates correspond to the start of week 1 and the end of week 43 in 2020 and 2019 respectively using a standard week numbering system (ISO 8601).^9^ Data prior to 1 April 2020 was taken from ‘annual refresh’ HES datasets, while data from 1 April 2020 was taken from a monthly extract to end October 2020. All datasets were supplied by NHS Digital under agreement DARS-NIC-194629-S4F9X.

We included activity data from all providers: English NHS hospitals, and English NHS commissioned activity in the independent sector.

### Outcomes and comparisons

We counted the total number of attended appointments (referred to in this paper as attendances, and covering both face-to-face and remote consultations) during each week in 2020, and compared against the equivalent weeks in 2019.

For the period between 2 March and 25 October 2020 (weeks 10 to 43 inclusive), we recorded the total cumulative difference in activity, the maximum weekly percentage difference, and the weekly percentage difference at the end of the period.

To allow us to make more appropriate comparisons, we adjusted the weekly counts of attendances to take account of public holidays, multiplying by 5/4 for weeks containing public holidays (note that the overwhelming majority of outpatient attendances in England tend to take place during the working week).

For sub-national analyses – by region of provider, and by groupings of patient characteristics (age and sex bands, ethnic groups, and deciles of socioeconomic deprivation assigned via Index of Multiple Deprivation 2019 linked at the lower super output area of residence) – we compared the 2020 adjusted weekly counts of attendances with week 9 values (24 February to 1 March 2020). Week 9 was selected to represent a period before any Covid-19-related impacts had become evident.

We documented adjusted weekly counts of attendances that took place remotely (by telephone or telemedicine) versus face-to-face, also calculating weekly ratios and comparing with weekly 2019 ratios. We additionally calculated weekly ratios of remote versus face-to-face attendances in 2020 for the aforementioned patient characteristic groupings.

We also carried out two analyses by clinical treatment specialty. Selecting the 30 largest clinical specialties as measured by activity at week 9 (accounting for over 80% of national outpatient activity that week), we firstly calculated adjusted weekly numbers of attendances by specialty, relative to week 9 values. Secondly, we calculated weekly ratios of face-to-face versus remote consultations by specialty.

The relative size of all analysis groups can be found in Appendix Table A1.

### Patient and public involvement

Two patient representatives were involved in reviewing a draft of the wider project of which this study was one component, but they were not directly involved in the design or implementation of the study. No patient representatives were asked to advise on interpretation or writing of the manuscript.

## RESULTS

### Total attendances

The outbreak of Covid-19 was associated with a large drop in outpatient attendances in England (Figure 1). Attendances began to diverge from 2019 levels at the beginning of March 2020, and fell rapidly following the publication of NHS guidance recommending that trusts free up inpatient capacity and postpone planned operations.

**Figure 1.**
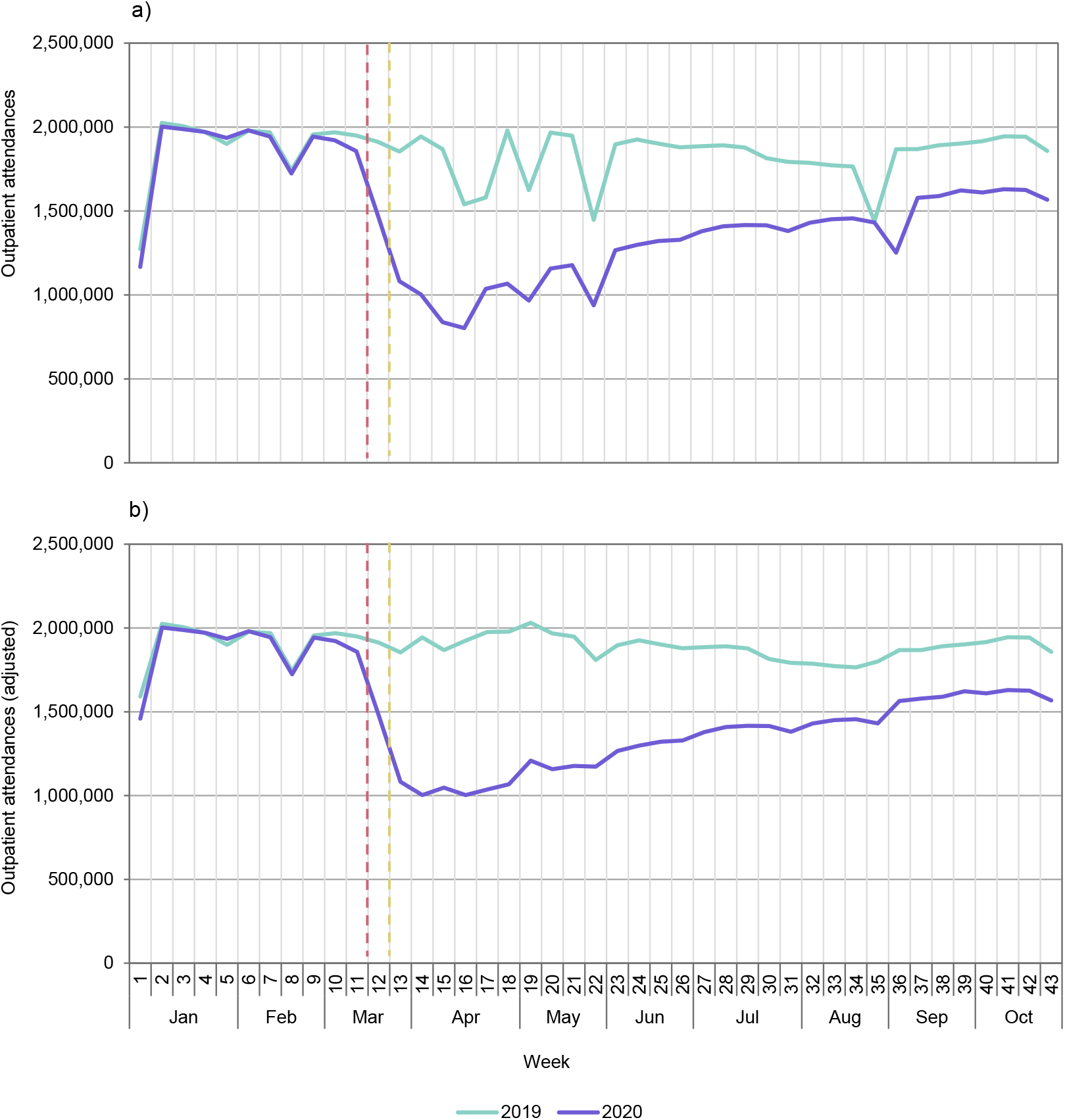
Weekly counts of outpatient attendances in 2019 and 2020 (to week 43) a) unadjusted figures b) adjusted for public holidays. Dashed lines indicate weeks in 2020 corresponding to NHS England COVID-19 letter (left) and start of national lockdown (right).

Between weeks 10 (beginning 2 March) and 43 (beginning 19 October), there was a total reduction of 16.6 million attendances compared to the same period in 2019. This is equivalent to 27% of outpatient attendances in these weeks in 2019. The largest weekly drop relative to 2019 (adjusted figures) was 48% in week 14 (week beginning 30 March). Subsequently, numbers of attendances increased but at a much slower rate than they fell; by week 43 they were still 16% lower than the equivalent week in 2019.

### Attendances by region

There was little difference in the timing of the fall in attendances for different NHS regions, with the bulk occurring in weeks 12 and 13 (16 to 29 March) (Figure 2). There were, however, some differences in the relative magnitude of reductions by region. For example, compared to week 9, the largest percentage drop in attendances occurred in the West Midlands (54%), while the smallest drop occurred in the South East (45%). By week 43, the number of attendances in all regions remained between 15% and 23% below the number in week 9.

**Figure 2.**
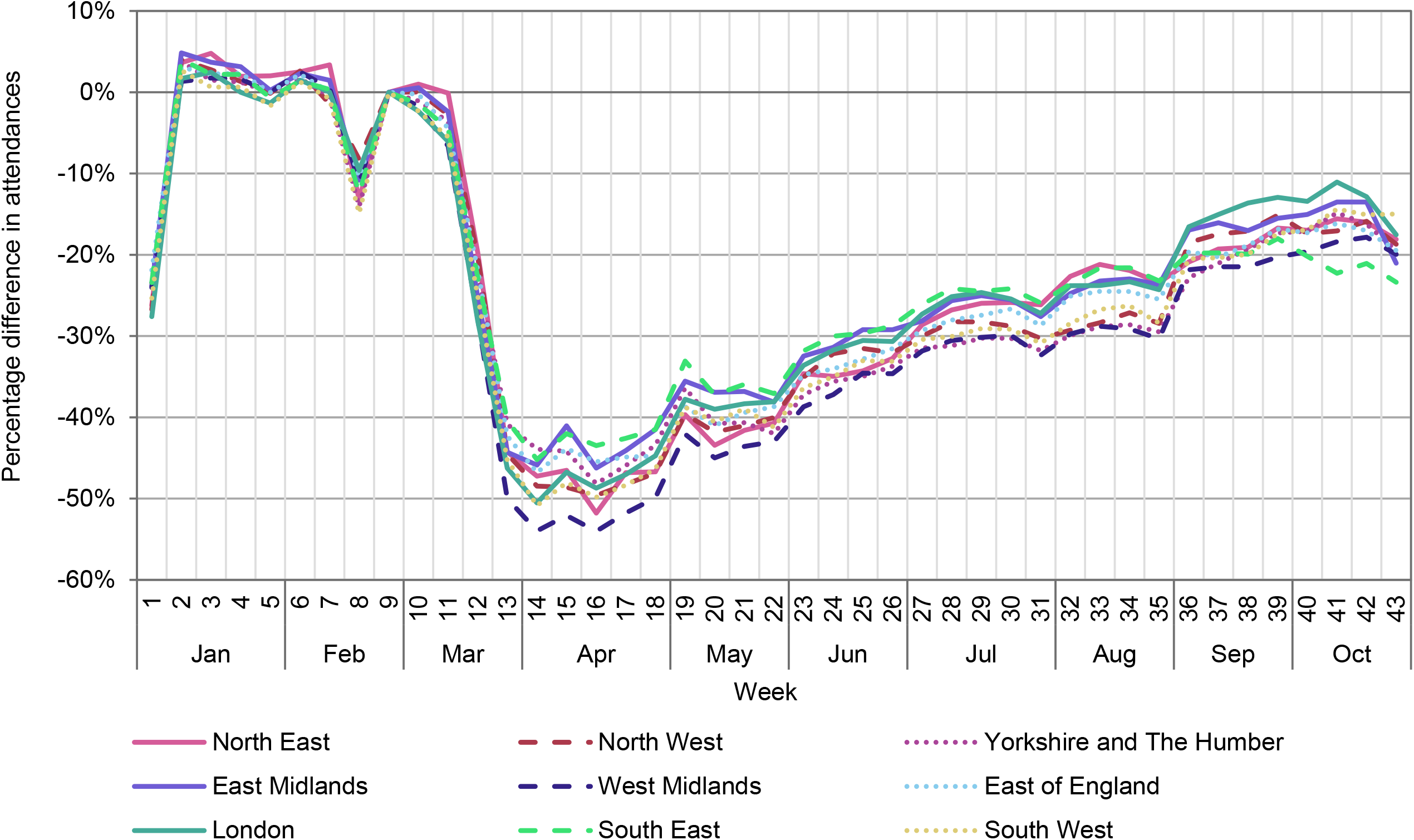
Weekly outpatient attendances (adjusted) by NHS region in 2020, (percentage change relative to week 9)

### Attendances by patient characteristics

Patterns of attendances were broadly similar across most patient characteristics: by age and sex, ethnic groups, and areas grouped by deciles of socioeconomic deprivation. The exceptions were in the case of females aged 18 to 39, and the relatively small mixed ethnic group, where the relative reductions in attendances after week 9 were less pronounced than for other groups (Figures 3, 4 and 5).

**Figure 3.**
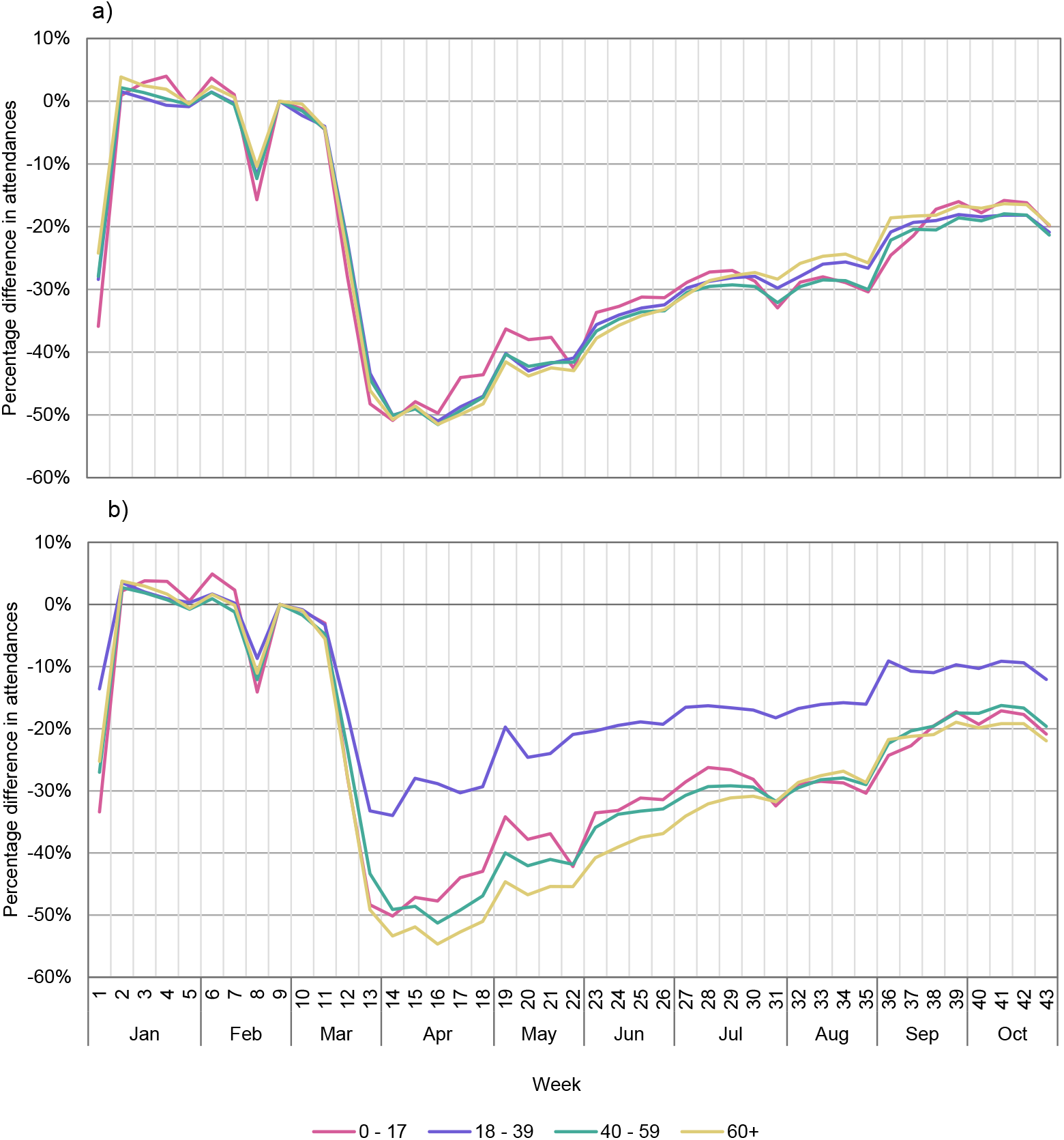
Weekly outpatient attendances (adjusted) by age and sex in 2020, (percentage change relative to week 9) a) males and b) females

**Figure 4.**
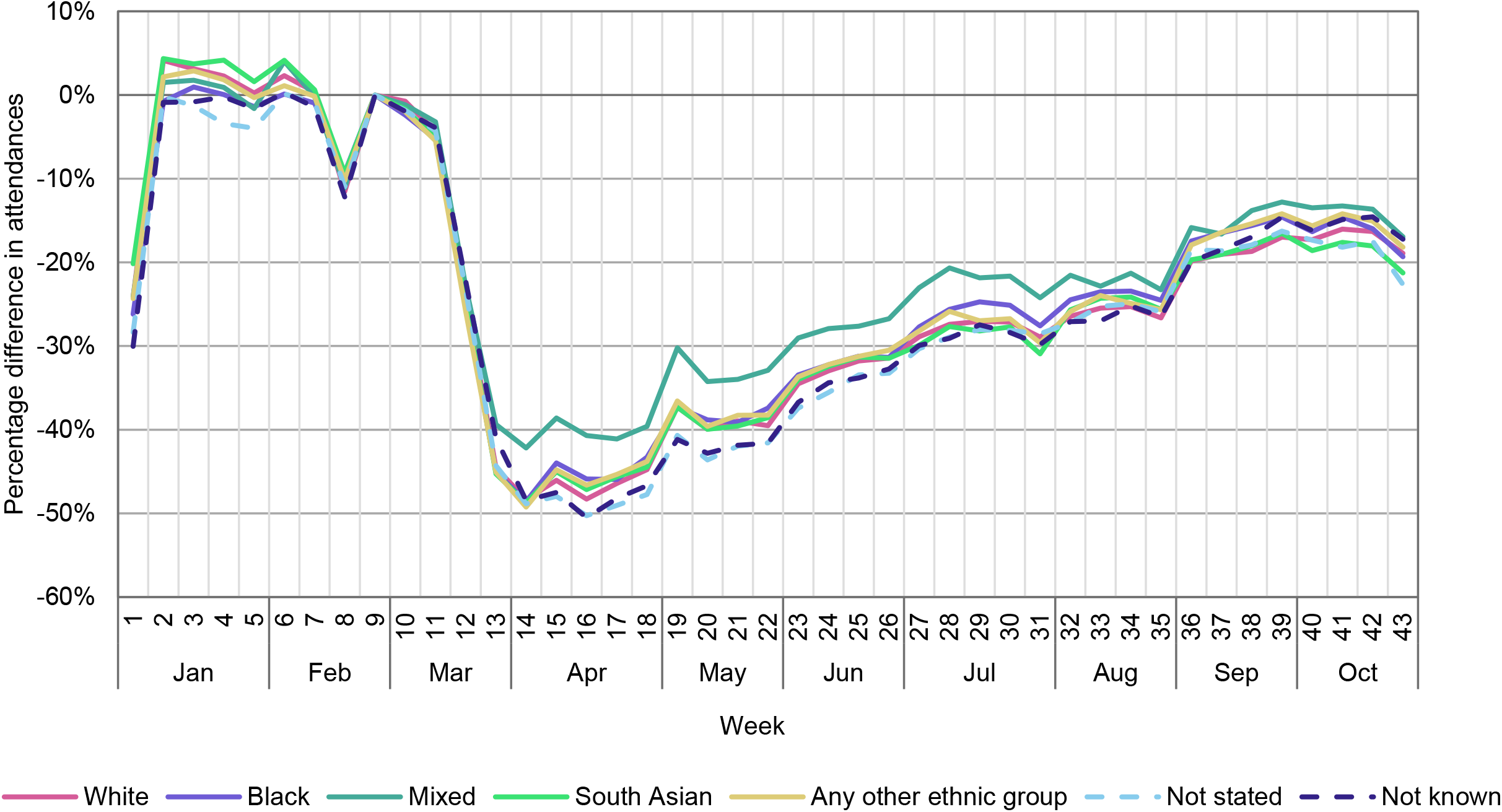
Weekly outpatient attendances (adjusted) by ethnicity group in 2020 (percentage change relative to week 9)

**Figure 5.**
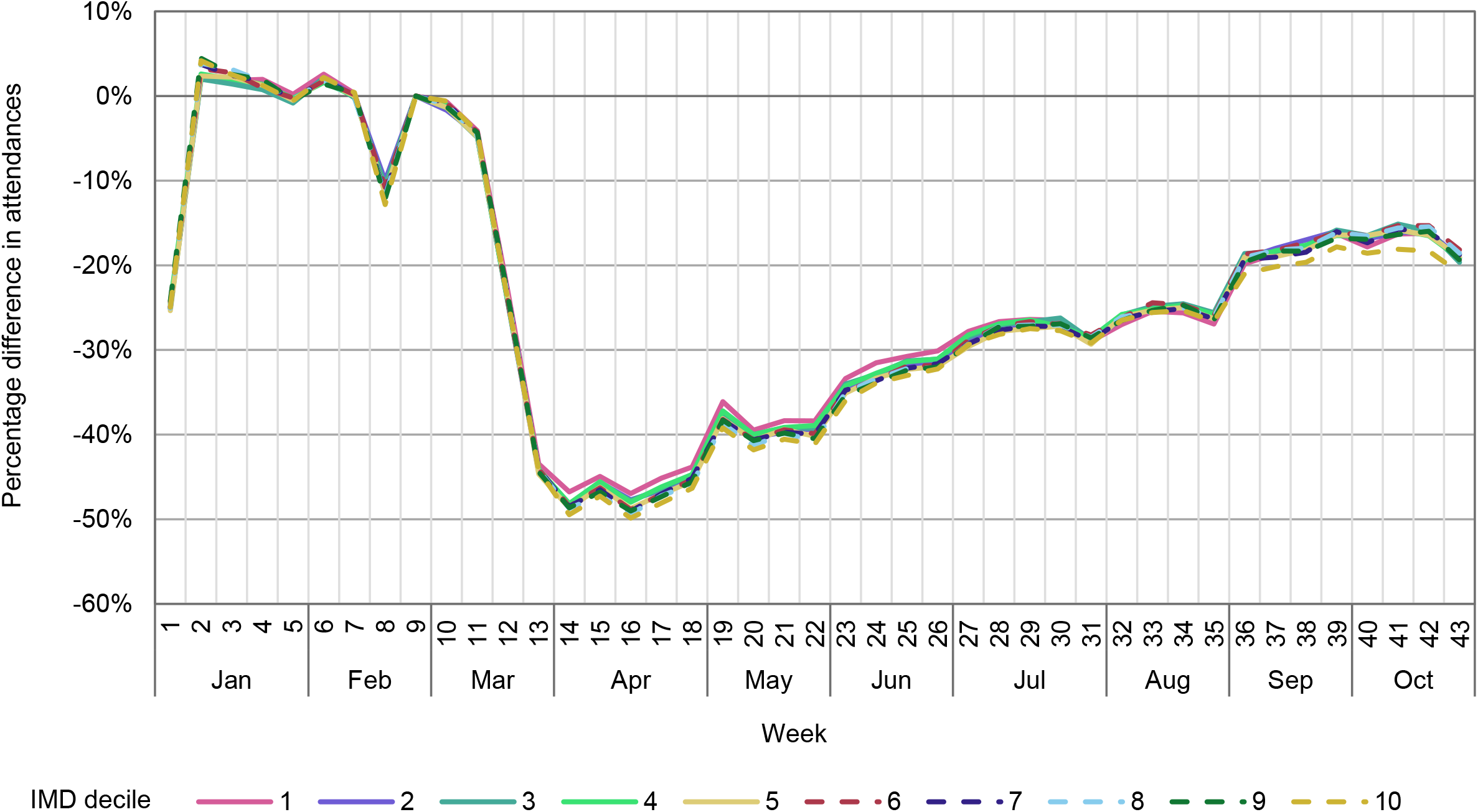
Weekly outpatient attendances (adjusted) by decile of socioeconomic deprivation in 2020 (percentage change relative to week 9), IMD decile 1 = most deprived and IMD decile 10 = least deprived

### Outpatient attendances carried out remotely

There was a significant increase in the proportion of consultations taking place remotely associated with the pandemic in 2020. Remote consultations increased from 4% of all outpatient attendances in week 9 (also 4% in the weeks before, including throughout 2019) to a high of 36% in week 18 (beginning 27 April 2020) (Figure 6). Subsequently, the proportion of remote consultations fell, although by week 43 they still represented 25% of all outpatient attendances. The relative reduction in remote attendances after week 18 was largely the result of an increase in face-to-face attendances, rather than a decline in the absolute number of remote attendances.

**Figure 6.**
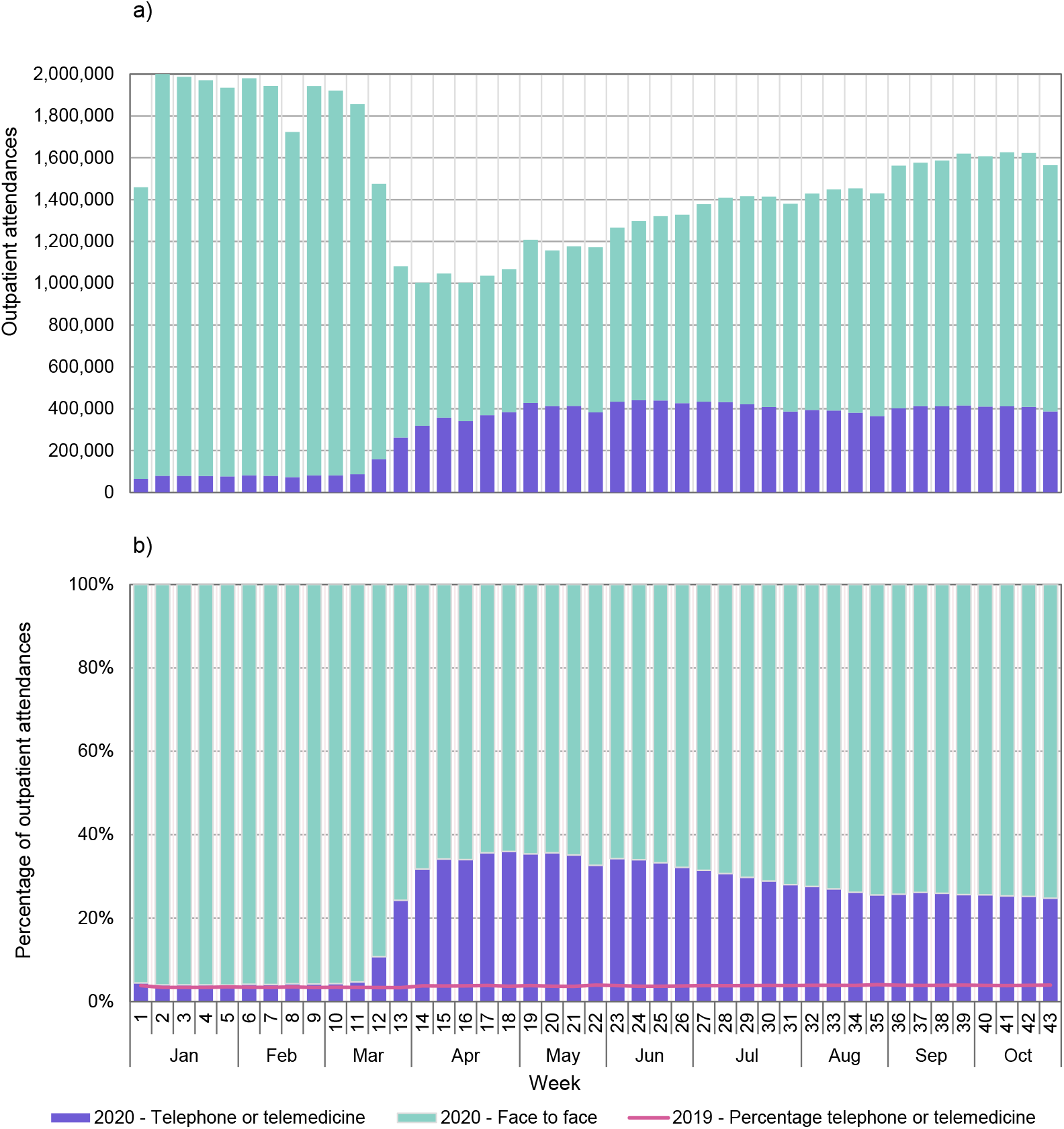
Mode of outpatient attendance: remote and face-to-face. A) Weekly adjusted counts of attendances in 2020, b) Weekly ratios of counts in 2020, and 2019 (line)

### Attendances carried out remotely by patient characteristics

Patterns of remote consultations were broadly similar for those in the 40-59 and 60 plus age groups, as well as for males aged 18-39. For males and females aged under 18, the proportion of remote attendances peaked at a higher level than for other age groups and then fell more rapidly. For women aged 18-39, remote attendances were markedly less common than for other groups (Figure 7).

**Figure 7.**
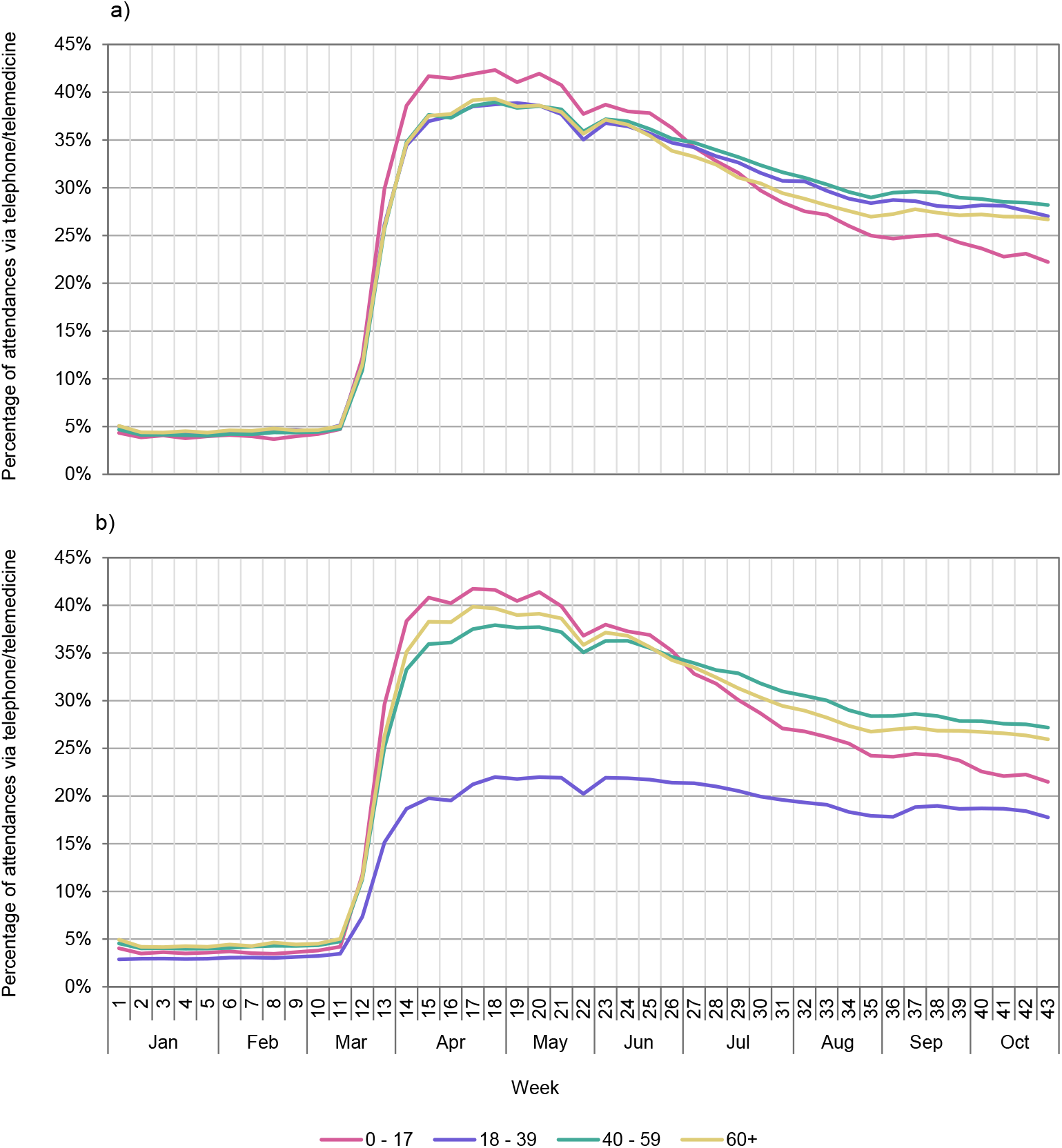
Weekly attendances held via remote means, 2020, by sex and age band. A) Males, B) Females

There was an apparent gradient by decile of socioeconomic deprivation, with patients in the most deprived areas tending to have lower rates of remote consultations than those in the least deprived areas (Figure 8). There was a noticeable difference between ethnic groups, with higher rates of remote attendances among white patients and lower rates among black, south Asian and other ethnic groups (Figures 9).

**Figure 8.**
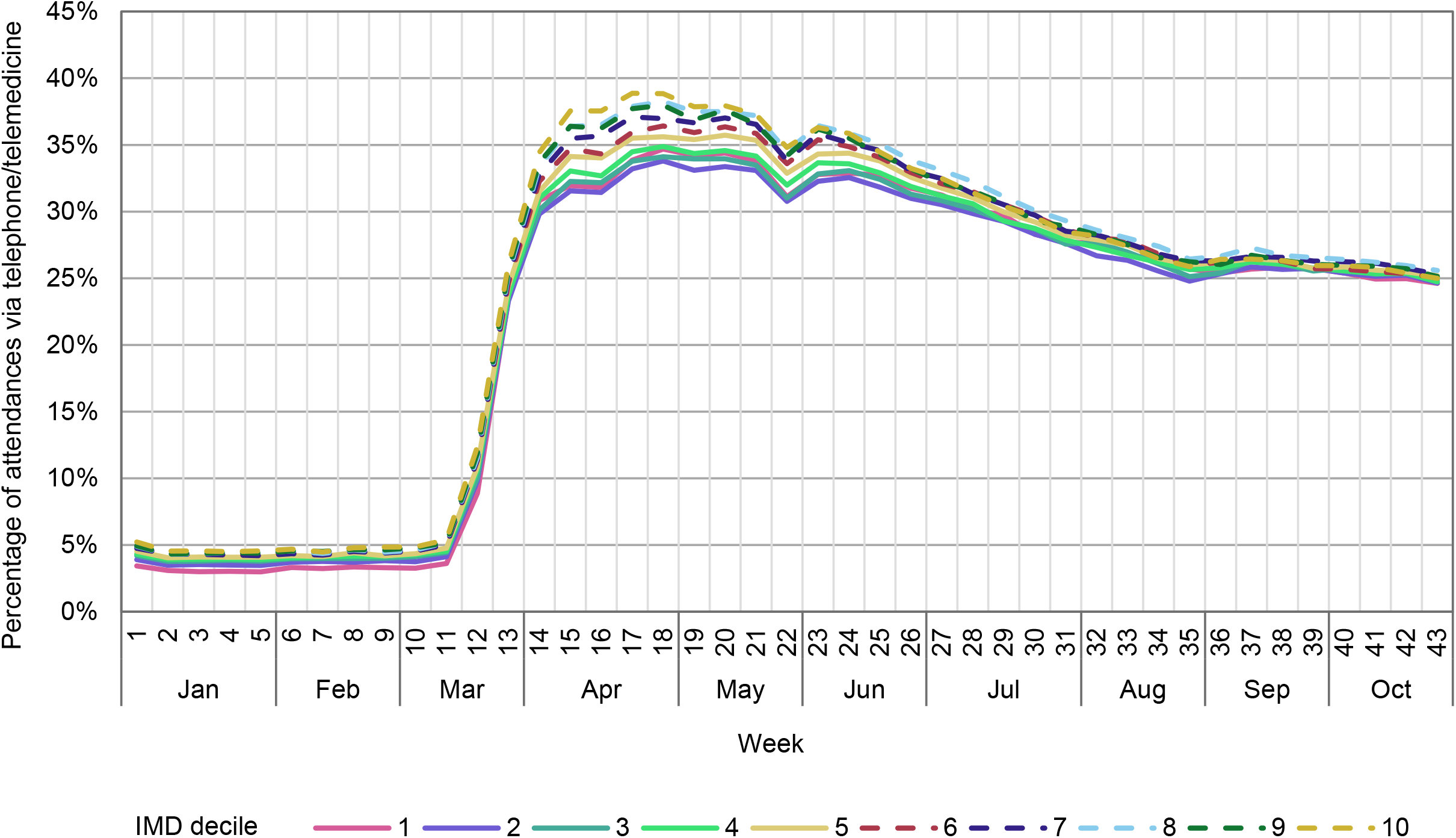
Weekly attendances held via remote means, 2020, by decile of deprivation. IMD decile 1 = most deprived and IMD decile 10 = least deprived.

**Figure 9.**
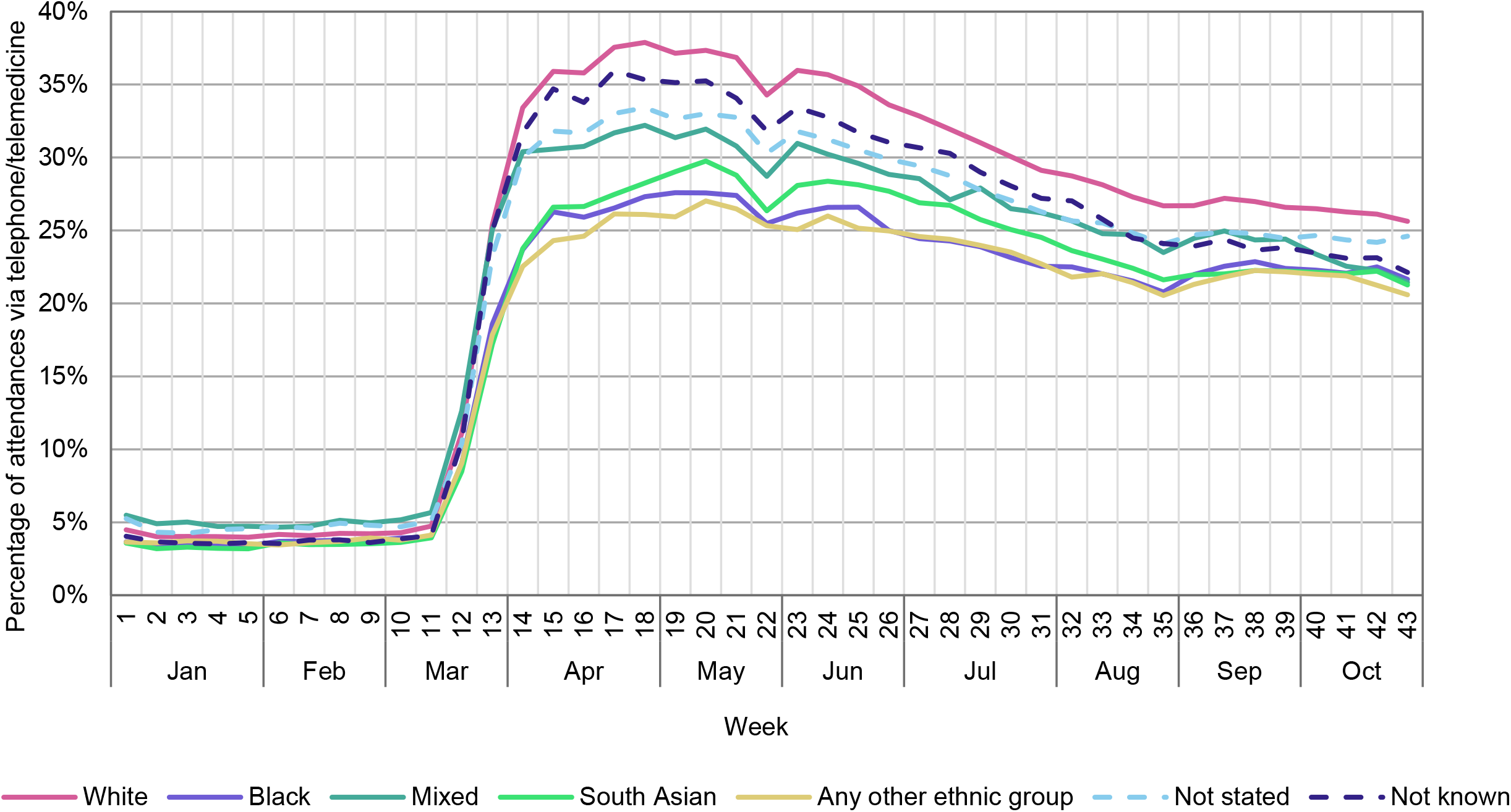
Weekly attendances held via remote means, 2020, by ethnic group.

### Attendances by clinical specialty

Trends in numbers of attendances varied considerably by clinical specialty (Figure 10). For example, attendances for audiology fell by 78% in week 17 compared to week 9, while those for medical oncology – the specialty with the most modest fall – fell by 11% (in week 13). Obstetrics, midwifery and clinical oncology were other services that showed relatively little variation over time. Note that for these specialities, the adjusted figures likely over-adjust for weeks containing public holidays (hence the peaks).

**Figure 10.**
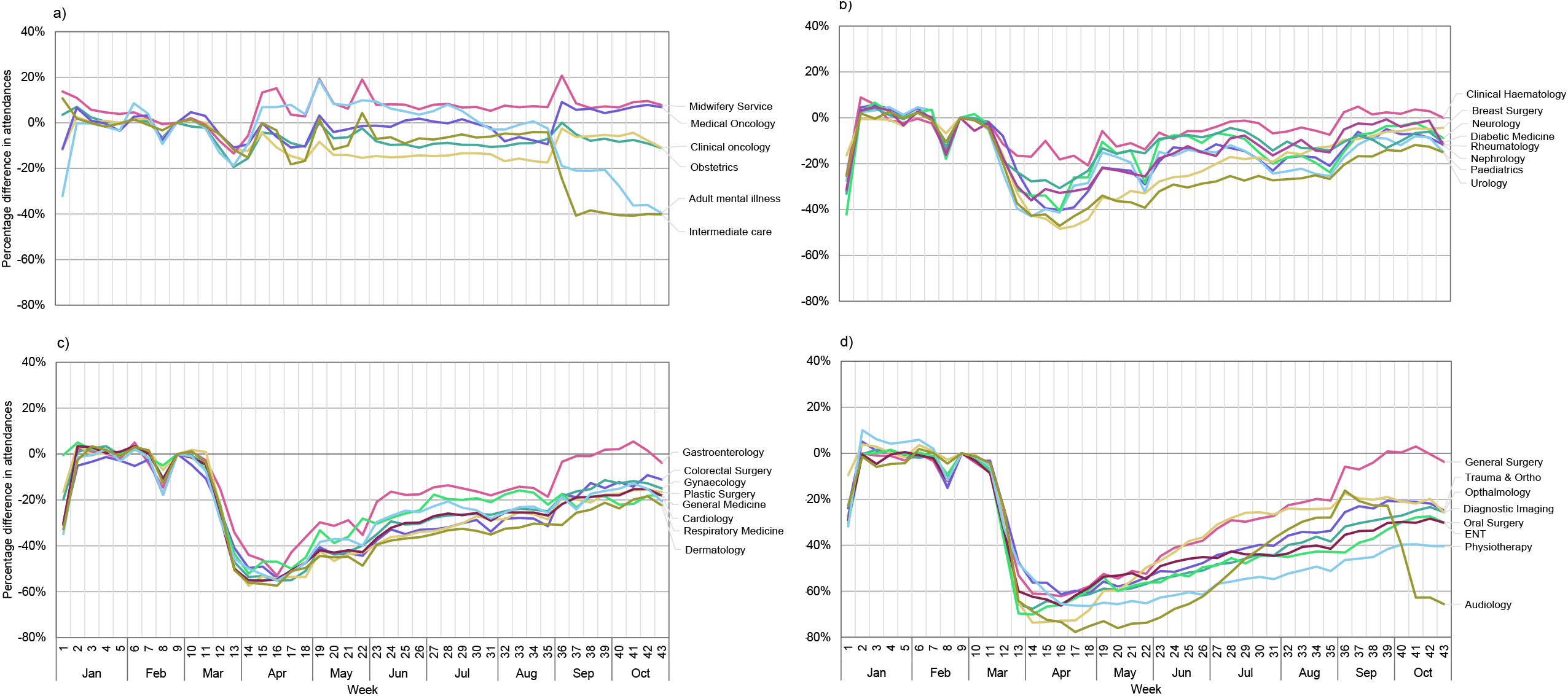
Weekly outpatient attendances (adjusted) by clinical specialty (percentage change relative to week 9). For convenience, displayed separately by groups of maximum weekly fall (between weeks 10 and 20) of a) <20% b) 20 to 50% c) 50 to 60% d) > 60%

The specialties also displayed differences in levels of recovery (in terms of numbers of attendances) by week 43. For example, general surgery (which saw a maximum drop of 62% in week 16) had recovered to a 4% drop (relative to week 9) by week 43. Whereas physiotherapy (which saw a maximum drop of 66% in week 18) was still 40% lower than week 9 values at week 43.

Three of the smaller specialities showed a rather erratic pattern of attendances.

### Attendances carried out remotely by clinical specialty

The proportion of attendances that were carried out remotely by telephone or video increased across nearly all clinical specialties from the middle of March 2020, and there were marked differences in the extent to which different specialties switched to remote care (Figure 11). For example, more than 60% of weekly gastroenterology consultations took place remotely at peak (around week 22; up from 14% in week 9), while midwifery services increased to 7% remote consultations at peak (weeks 17 to 24; up from 1% in week 9). Diagnostic imaging consultations remained overwhelmingly face-to-face.

**Figure 11.**
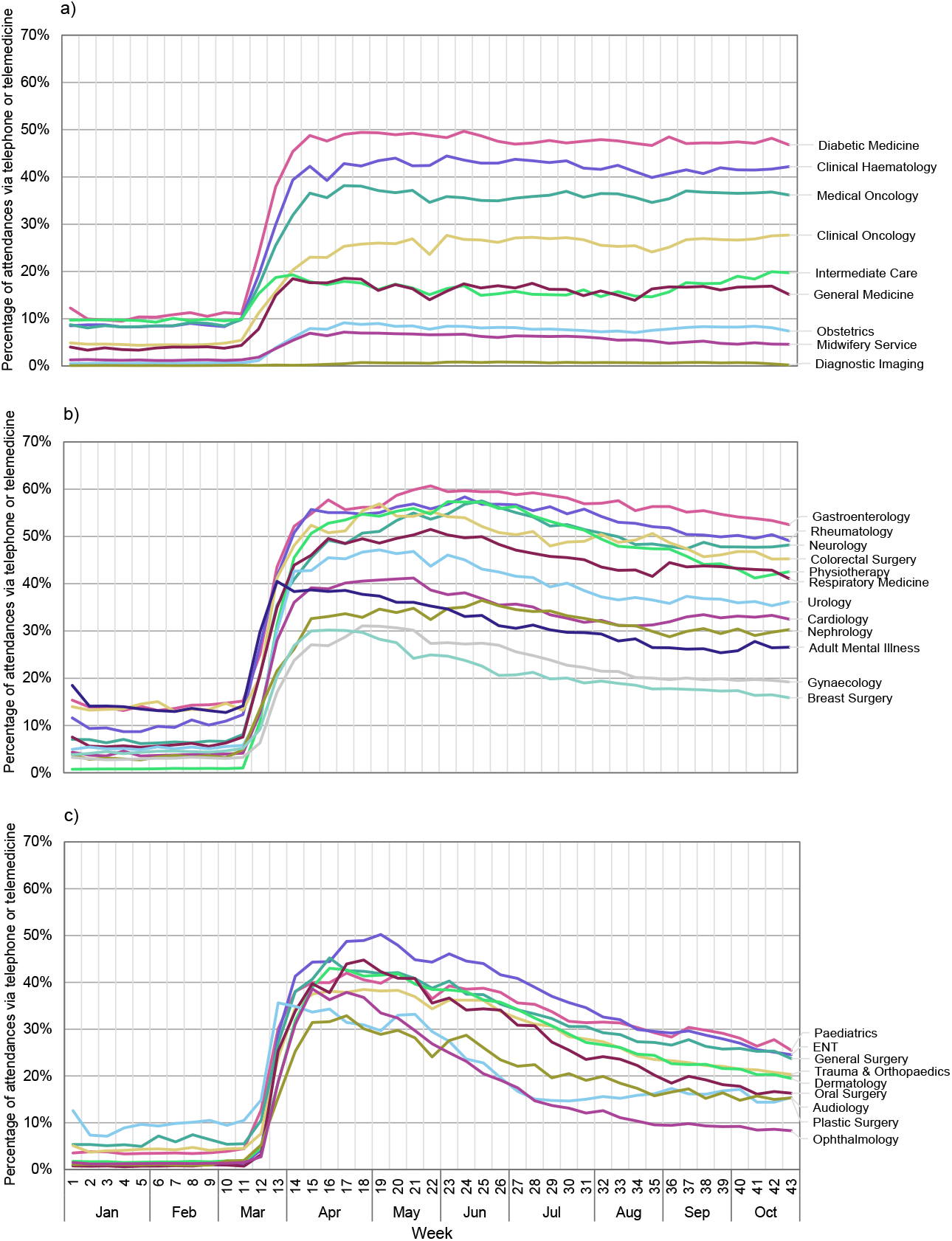
Percentage of weekly outpatient attendances (adjusted) carried out remotely by clinical specialty. For convenience, displayed separately by groups of maximum percentage point (ppt) difference from peak to week 43 a) <5 ppt b) 5 to 15 ppt c) >15 ppt

A further pattern of interest between specialties lay in the extent to which increases in remote attendances from March 2020 persisted across the rest of the year. A number of specialties (including diabetic medicine, medical and clinical oncology, and clinical haematology) broadly held steady over the period in their relative use of remote versus face-to-face consultations (Figure 11a); others (including ophthalmology, oral surgery and ear, nose and throat) had by week 43 fallen by up to 30 percentage points from their peak remote care proportion (Figure 11c).

## DISCUSSION

We report on a substantial decline of more than one quarter of outpatient attendances in England following the outbreak of the Covid-19 pandemic. The trend was one of an initial large fall between March and April 2020, followed by a gradual, although far from complete, recovery in numbers to late October 2020. We found almost no difference in the timing of the initial decline across NHS regions, and relatively little difference in changes in attendances across sex, age, ethnicity and proxy deprivation groups. We also found a very large increase in attendances that took place remotely; more than one in three were remote at peak (during April and May 2020) falling to one in four by late October. We observed some variation in the use of remote care by demographic groups of patients; patients living in more deprived areas had lower rates of remote consultations, and the same was true for ethnic minority patients (specifically Black, South Asian and Other ethnic groups). Finally, clinical specialties showed differing patterns in how their activity levels changed and in the extent to which they switched to remote consultations.

A major strength of the study is its nationwide population-based approach, which included all recorded outpatient activity in England’s NHS hospitals, in addition to NHS commissioned activity in independent providers. A disadvantage is that the analysis is largely descriptive and does not allow us to draw conclusions about, for example, the impact on the quality of outpatient care, or on the health and experience of patients. While the data was up-to-date at the time of analysis, trends in outpatient activity have continued to change. No formal statistical tests of differences were carried out (however, the numbers of events were relatively large) and adjustments for public holidays were crude. Furthermore, some of the differences we observed between groups of patients (for example, ethnic groups) may be partly explained by differences in the demographic structure and relative prevalence of conditions in these groups; our analysis did not account for these factors. It is also possible that the accuracy of the data may have been affected by the pandemic in ways we do not understand. For example, clinical coding of remote versus face-to-face appointments may not have been consistent across services or over time.

Published data shows that the trends observed in outpatient attendances mirrored similar changes in activity across other hospital services in England, including elective and emergency admissions, and attendances at emergency departments.^10,11^ A similar trend in outpatients was also observed in the United States, where attendances fell by almost 60% in April 2020 but had largely recovered by October.^12^ The lack of variation in the timing of the initial decline in attendances across NHS regions does not match the geographical differences in Covid-19 case numbers and death rates at the outset of the pandemic.^13^ This suggests that the initial reductions were related to hospitals across the country anticipating an impending wave of virus impacts and responding to instructions to free-up inpatient care, rather than being direct effects of local levels of illness.

An exception to the lack of variation in changes in attendances between demographic groups was for women aged 18 to 39 for whom falls in attendances were more modest than other groups. This is likely to be connected to the observation that activity levels for midwifery and obstetrics specialties continued throughout the period at broadly pre-pandemic levels. Attendances also remained largely stable for medical oncology, clinical oncology and clinical haematology – probably a reflection of the services’ prioritisation or ringfencing of activity for more urgent and time-critical cases. In comparison, clinical specialties such as ophthalmology, physiotherapy and audiology exhibited very large declines of 60-80% at their minimum points. This may partly reflect the postponement of less urgent cases but also a lack of suitability of remote consultation for certain clinical specialties.

We note that the proportion of attendances that took place remotely in October is in line with NHS England’s 2021/22 planning guidance, which recommends that where attendances are clinically necessary, at least 25% should be delivered remotely.^14^ It remains to be seen to what extent this is sustained. More detailed work is also needed to understand whether variations in remote care use between demographic groups of patients reflect significant differences in healthcare access.

Further work is needed to better understand the differing patterns in the extent to which clinical specialties switched to remote consultations, and the ability of some specialties to maintain high levels of remote care while others declined over time. Some variations may in part reflect the nature of the specialty – for example, the inability to carry out diagnostic imaging remotely. Also unknown is whether the switch to remote consultations has resulted in comparable levels of quality of care and patient outcomes; this remains an outstanding evaluative question.

## Conclusions

The Covid-19 pandemic has led to large-scale change in outpatient service provision. The main aim of this analysis was to describe changes in outpatient activity in England over the initial months of the pandemic in order to inform future work to identify and evaluate innovations in how outpatient services are delivered. The two major changes have been the overall drop in activity, and the rise in remote consultations. With respect to the latter there has been variation in patterns of remote consultation between patient groups. Along with impacts that remote care may be having on patient experiences and outcomes as well as on the efficiency with which the NHS delivers outpatient services, this highlights the need to evaluate the reasons for these differences.

## Data Availability

The authors do not have permission to share data with third parties, but the data may be accessed through NHS Digital via an appropriate agreement.

## ACKNOWLEDGEMENTS

We would like to thank NHS Digital staff for the provision of data for use by NIHR Rapid Service Evaluation Team (RSET). The Hospital Episode Statistics data are copyright NHS Digital. We would also like to thank our colleagues Sarah Reed and Jonathan Spencer for their advice.

## CONTRIBUTORS

JM, TG and JA (guarantor) contributed to the design of the study. JM was responsible for the data analysis. JM, TG and JA were all involved in the interpretation of data, contributing to drafts of the manuscript and reviewing the final version. The corresponding author attests that all listed authors meet authorship criteria and that no others meeting the criteria have been omitted.

## FUNDING

This work was funded by the National Institute for Health Research (NIHR) (Health Services and Delivery Research, 16/138/17 – Rapid Service Evaluation Team). The views expressed are those of the authors and not necessarily those of the NIHR or the Department of Health and Social Care.

## COMPETING INTERESTS

Competing interests: All authors have completed the ICMJE uniform disclosure form at www.icmje.org/coi_disclosure.pdf and declare: financial support from NIHR for the submitted work; no financial relationships with any organisations that might have an interest in the submitted work in the previous three years; no other relationships or activities that could appear to have influenced the submitted work.

## ETHICAL APPROVAL

Ethical approval was not needed for this study.

## DISSEMINATION TO PARTICIPANTS AND RELATED PUBLIC COMMUNITIES

The size of the study population rules out direct dissemination to participants.

**Appendix Table A1.**
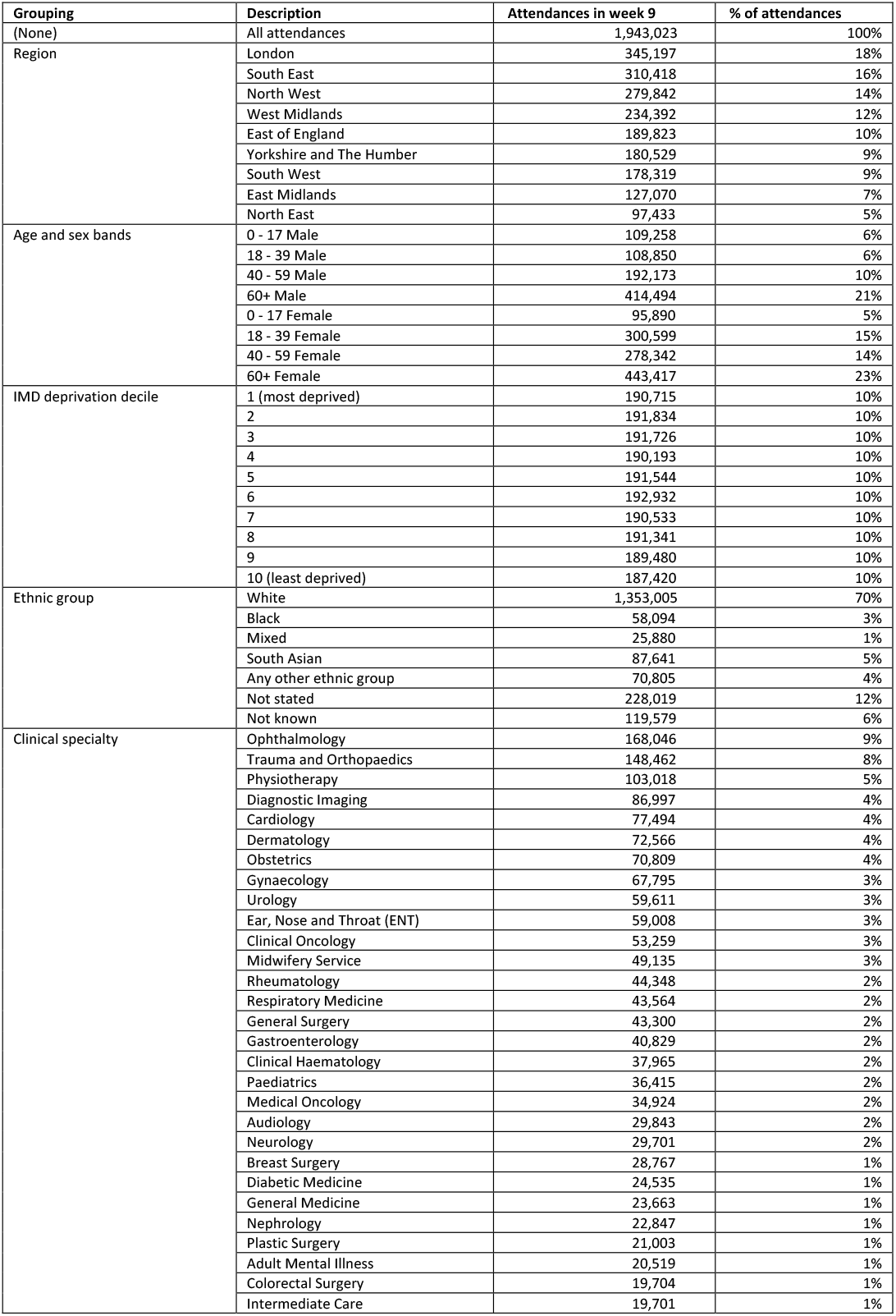
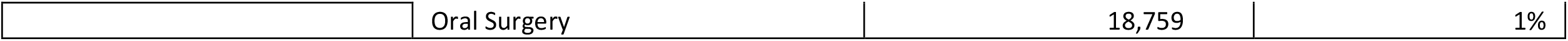
Size of groups (number of attendances in Week 9, 2020 – pre-COVID)

| Grouping | Description | Attendances in week 9 | % of attendances |
| --- | --- | --- | --- |
| (None) | All attendances | 1,943,023 | 100% |
| Region | London | 345,197 | 18% |
|  | South East | 310,418 | 16% |
|  | North West | 279,842 | 14% |
|  | West Midlands | 234,392 | 12% |
|  | East of England | 189,823 | 10% |
|  | Yorkshire and The Humber | 180,529 | 9% |
|  | South West | 178,319 | 9% |
|  | East Midlands | 127,070 | 7% |
|  | North East | 97,433 | 5% |
| Age and sex bands | 0 - 17 Male | 109,258 | 6% |
|  | 18 - 39 Male | 108,850 | 6% |
|  | 40 - 59 Male | 192,173 | 10% |
|  | 60+ Male | 414,494 | 21% |
|  | 0 - 17 Female | 95,890 | 5% |
|  | 18 - 39 Female | 300,599 | 15% |
|  | 40 - 59 Female | 278,342 | 14% |
|  | 60+ Female | 443,417 | 23% |
| IMD deprivation decile | 1 (most deprived) | 190,715 | 10% |
|  | 2 | 191,834 | 10% |
|  | 3 | 191,726 | 10% |
|  | 4 | 190,193 | 10% |
|  | 5 | 191,544 | 10% |
|  | 6 | 192,932 | 10% |
|  | 7 | 190,533 | 10% |
|  | 8 | 191,341 | 10% |
|  | 9 | 189,480 | 10% |
|  | 10 (least deprived) | 187,420 | 10% |
| Ethnic group | White | 1,353,005 | 70% |
|  | Black | 58,094 | 3% |
|  | Mixed | 25,880 | 1% |
|  | South Asian | 87,641 | 5% |
|  | Any other ethnic group | 70,805 | 4% |
|  | Not stated | 228,019 | 12% |
|  | Not known | 119,579 | 6% |
| Clinical specialty | Ophthalmology | 168,046 | 9% |
|  | Trauma and Orthopaedics | 148,462 | 8% |
|  | Physiotherapy | 103,018 | 5% |
|  | Diagnostic Imaging | 86,997 | 4% |
|  | Cardiology | 77,494 | 4% |
|  | Dermatology | 72,566 | 4% |
|  | Obstetrics | 70,809 | 4% |
|  | Gynaecology | 67,795 | 3% |
|  | Urology | 59,611 | 3% |
|  | Ear, Nose and Throat (ENT) | 59,008 | 3% |
|  | Clinical Oncology | 53,259 | 3% |
|  | Midwifery Service | 49,135 | 3% |
|  | Rheumatology | 44,348 | 2% |
|  | Respiratory Medicine | 43,564 | 2% |
|  | General Surgery | 43,300 | 2% |
|  | Gastroenterology | 40,829 | 2% |
|  | Clinical Haematology | 37,965 | 2% |
|  | Paediatrics | 36,415 | 2% |
|  | Medical Oncology | 34,924 | 2% |
|  | Audiology | 29,843 | 2% |
|  | Neurology | 29,701 | 2% |
|  | Breast Surgery | 28,767 | 1% |
|  | Diabetic Medicine | 24,535 | 1% |
|  | General Medicine | 23,663 | 1% |
|  | Nephrology | 22,847 | 1% |
|  | Plastic Surgery | 21,003 | 1% |
|  | Adult Mental Illness | 20,519 | 1% |
|  | Colorectal Surgery | 19,704 | 1% |
|  | Intermediate Care | 19,701 | 1% |
|  | Oral Surgery | 18,759 | 1% |

## STROBE Statement

Checklist of items that should be included in reports of observational studies

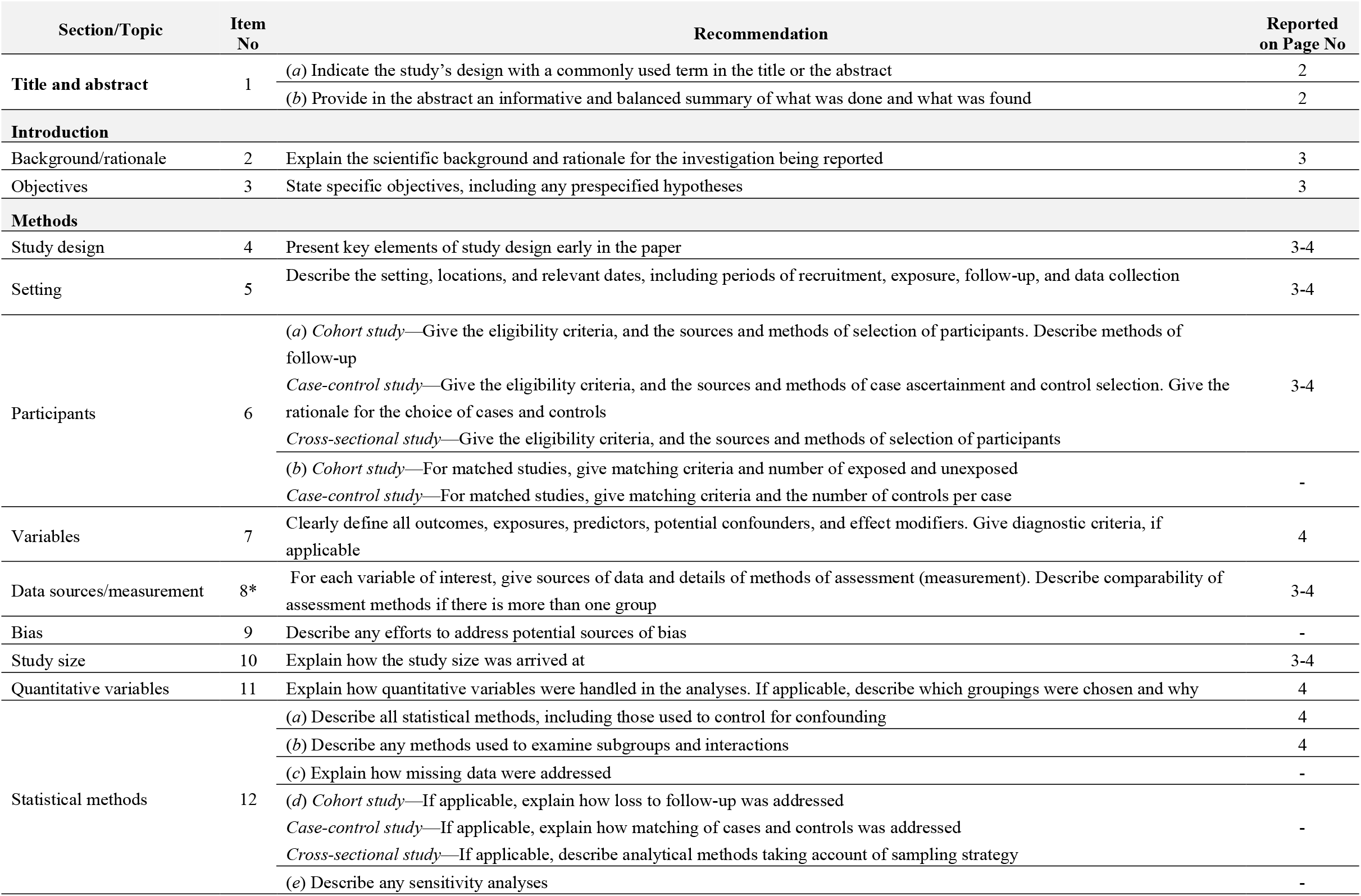
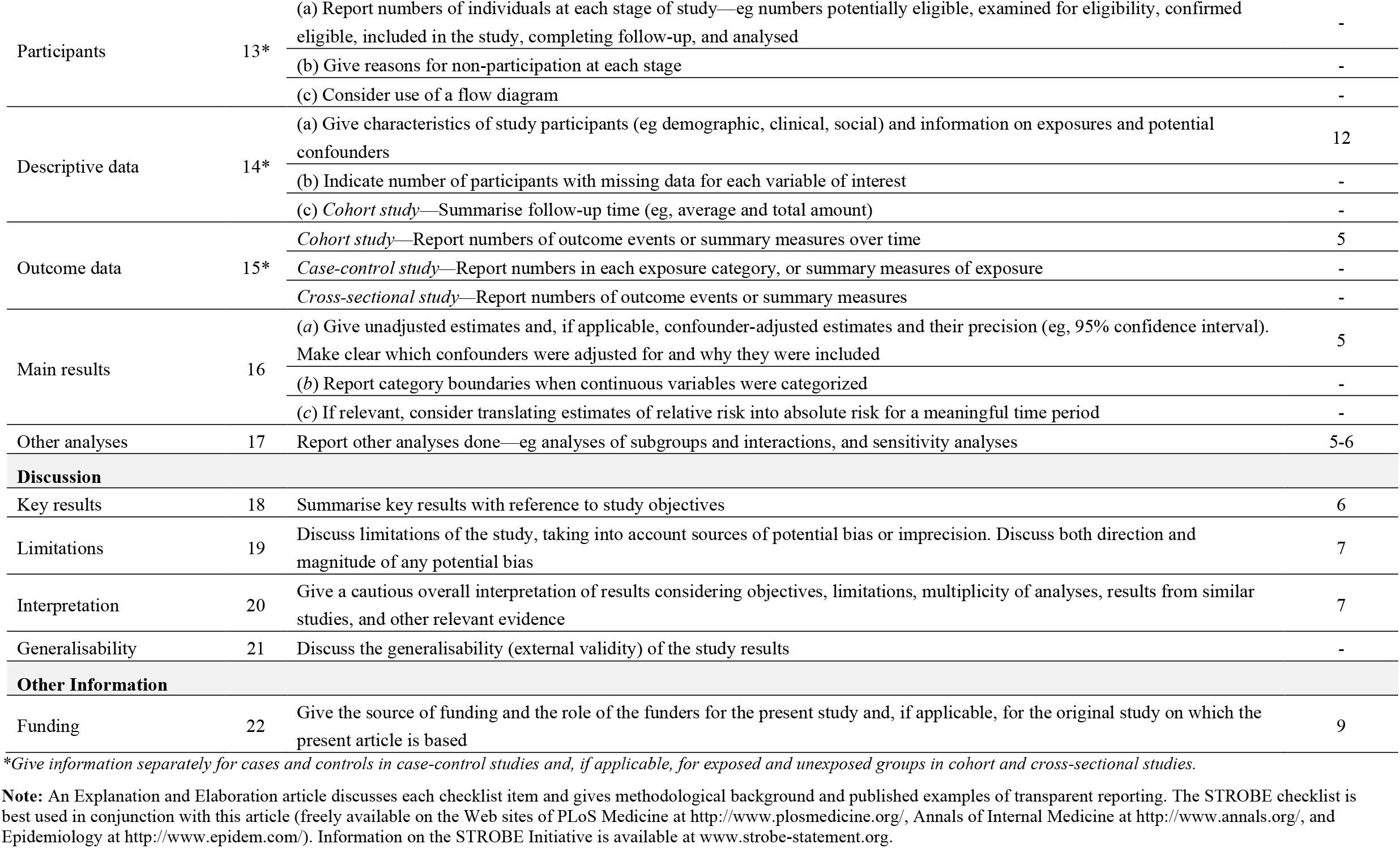

## Notes

### Funding Statement

This work was funded by the National Institute for Health Research (NIHR) as part of its NIHR Rapid Service Evaluation Team (RSET), award ID 16/138/17. The funder had no role in the study design or in the collection, analysis, interpretation of data, writing of the report, or decision to submit the article for publication.

### Author Declarations

Ethical approval was not needed for this study.

